# Severe and uncomplicated *Plasmodium knowlesi* malaria in North Kalimantan and Sabang, Aceh, and suspected *Anopheles* vectors across Kalimantan, Indonesia

**DOI:** 10.1101/2025.04.29.25326532

**Authors:** Leily Trianty, Ristya Amalia, Rintis Noviyanti, Pinkan Kariodimedjo, Nadia Fadila, Fahira Ainun Nisa, Farah Novita Coutrier, Titik Yuniarti, Irnawati Irnawati, Hidar, Thomas R Burkot, Kim A Piera, Bridget E Barber, Nicholas M Anstey, Triwibowo A Garjito, Matthew J Grigg

## Abstract

**Background:** *Plasmodium knowlesi* is an emerging cause of zoonotic malaria in Southeast Asia. Current detection tools limit insight into the extent of transmission across Indonesia. This study aimed to enhance molecular and entomological surveillance around clinical cases to evaluate *P. knowlesi* transmission in Kalimantan and Aceh, Indonesia.

**Methods:** Febrile patients presenting to district-level health facilities in Malinau, North Kalimantan, and Sabang, Aceh, were screened for malaria using routine diagnostic tools and ultrasensitive reverse-transcriptase PCR. Clinical and epidemiological data were collected. Overnight human landing catch mosquito surveys were conducted around *P. knowlesi* case locations in North Kalimantan. Anophelines were morphologically identified and compared to 2016-18 Ministry of Health entomology data across four Kalimantan provinces. Generalised linear mixed models (GLMM) compared mosquito group distributions across habitats.

**Results:** From December 2019 to September 2020, 429 patients were enrolled. *Plasmodium knowlesi* was the major cause of malaria transmission in Malinau and Sabang. In Malinau, 15/81 (18.5%) patients had malaria, comprising 10 (12.2%) *P. knowlesi* and 5 (6.1%) *P. vivax* infections. In Sabang, 6/348 (1.7%) patients had malaria, including 4 (1.1%) *P. knowlesi* infections. Two patients from Malinau met WHO-criteria for severe disease (hyperparasitaemia >100,000/µL). Microscopy and pan-pLDH-based rapid diagnostic tests had >90% sensitivity for detecting *P. knowlesi*, although misidentification with *P. vivax* occurred. Risk-associations with *P. knowlesi* infection included forest-related activities, with all cases in adult males. *Anopheles leucosphyrus* group vectors (*An. balabacensis* and *An. latens*) were identified in farm, forest, and village environments in North Kalimantan, in coastal (*An. hackeri*), farm and forest (*An. leucosphyrus*) sites in Central Kalimantan, and forest sites (*An. leucosphyrus*) in South Kalimantan. GLMM analyses predicted fewer *Anopheles leucosphyrus* and *barbirostris* group mosquitoes in coastal habitats. *Anopheles* species diversity was highest in farm environments (mean Shannon-Weiner index H′=0.84, SD=0.49).

**Conclusions:** *Plasmodium knowlesi* is a common cause of febrile illness and severe malaria in North Kalimantan. Findings support the need for ongoing access to appropriate diagnostics and treatment in non-zoonotic malaria elimination-phase settings. Competent vectors were found beyond traditional forest habitats across Kalimantan, with targeted vector monitoring and strengthened molecular surveillance essential for zoonotic malaria control efforts in Indonesia.

## Introduction

More than 8% of the population in Indonesia live in areas of high malaria transmission, with *P. falciparum* infections in >62% of cases and the highest disease burden in the eastern province of Papua^1^. However, zoonotic malaria transmission with the macaque parasite *P. knowlesi* complicates recent national reporting estimates, with *P. knowlesi* human infections molecularly confirmed across large areas of western Indonesia^2^ including the provinces of Aceh^3–7^, North Sumatra^8^, Jambi^9^, Central Kalimantan^10,11^, East Kalimantan^12^ and South Kalimantan^13,14^. National WHO malaria certification has recently been provisionally modified to require negligible reporting (below 10 cases) of *P. knowlesi* each year^15^. Due to an absence of any systematic routine molecular testing in most at-risk areas in western Indonesia, a likely considerable underestimate of 87 local *P. knowlesi* cases were reported nationally in 2022^16^.

The widespread transmission of zoonotic malaria in Indonesia is consistent with regional confirmation of *P. knowlesi* cases and predictive risk mapping encompassing rapid agricultural land use change in areas where the long-tailed (*Macaca fasicularis*) and pig-tailed (*M. nemestrina*) macaques hosts and *Anopheles leucosphyrus* group vectors are present^17–19^. However, in the Indonesian studies to date, *P. knowlesi* has been universally misdiagnosed by microscopy as non-zoonotic *Plasmodium* species, with *P. falciparum* and *P. vivax* more commonly misidentified than the morphologically similar *P. malariae*^14,20^. The majority of earlier malaria lateral flow-based rapid diagnostic tests were insufficiently sensitive and specific for *P. knowlesi* detection at low parasite counts, with cross-reactivity occurring against the commonly used *P. vivax* and *P. falciparum* parasite lactate dehydrogenase targets^21–24^. On the island of Sabang, off the northern tip of Aceh Besar, *P. knowlesi* has been molecularly confirmed as the major cause of human malaria^4^ . In contrast, molecular surveillance for zoonotic malaria has not been conducted in North Kalimantan, despite reports of *P. knowlesi* in other Kalimantan provinces^2,11,14^ and the high number of reported cases of *P. knowlesi* malaria across the border in Sarawak and Sabah, Malaysia^32^.

Determining the presence and diversity of competent mosquito vectors is crucial to understanding spatial heterogeneity in zoonotic malaria transmission^25,26^. In parts of Kalimantan, *An. balabacensis* and *An. latens*, both members of the *An. leucosphyrus* group, have previously been identified^27,28^, with each confirmed or suspected vectors of zoonotic *P. knowlesi*^25,29–31^. Similarly, on Sabang Island, Aceh, at least 11 *Anopheles* species have been recorded^32^, including *An. dirus,* also a known member of the *An. leucosphyrus* group and a confirmed vector of *P. knowlesi* in Vietnam^33,34^ and suspected vector in neighbouring Peninsular Malaysia^31^. However, molecular testing of mosquito species in Sabang^32^, or other areas including Central Kalimantan^35^ have not detected *P. knowlesi*-specific DNA using well-established 18S rRNA gene targets to date. This finding suggests that either the *P. knowlesi* infection prevalence in these mosquitoes is low, sampling numbers are insufficient, or additional ecological factors influence vector competence in this region; highlighting the need for ongoing targeted entomological surveillance to assess the true risk of zoonotic malaria transmission.

This study aims to compare the human incidence and clinical and epidemiological features of zoonotic *Plasmodium* species infections between North Kalimantan and Sabang, Aceh, and to characterise the broader distribution of *Anopheles leucosphyrus* group vectors across Kalimantan.

## Methods

### Study sites

This health facility-based survey with passive malaria case detection was conducted in two distinct areas of western Indonesia: Malinau in North Kalimantan, and the island of Sabang, Aceh (**Figure 1**). There were five health facility study sites: a district referral hospital in Malinau and four health facilities across Sabang (Cot Bau, Sukajaya, Sukakarya, and Iboih). Field sampling of mosquito vectors was conducted through spot surveys in five villages of Malinau District with confirmed *P. knowlesi* cases: Malinau, Pulau Sapi, Long Metut, Long Tarik, and Jelukut.

**Figure 1:**
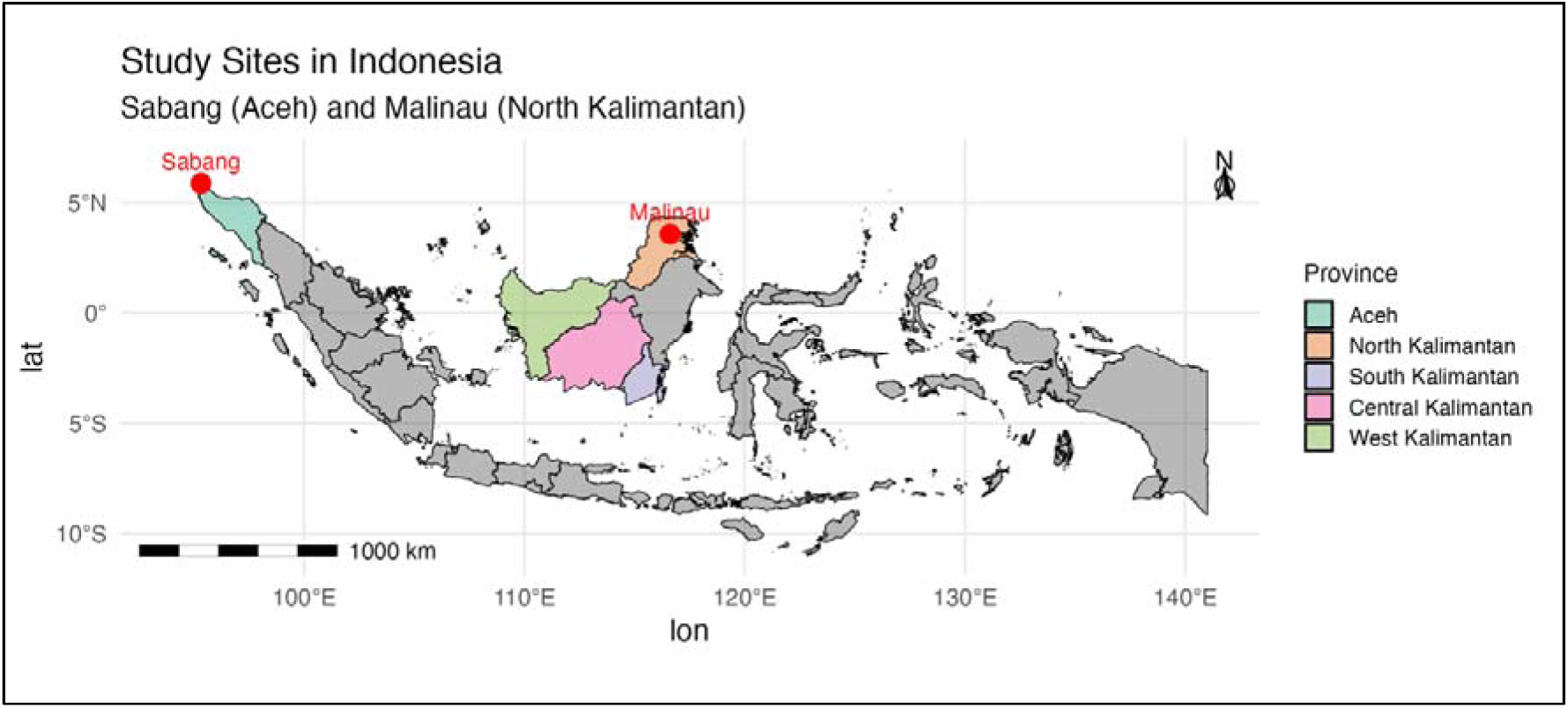
Map of study sites in Indonesia - Sabang, Aceh and Malinau, North Kalimantan (health facility surveys) and Kalimantan provinces (entomological surveillance)

### Study participants

Patients attending study health facilities were enrolled and screened for malaria if meeting the following inclusion criteria: age at least 1-year, documented body temperature of greater than 37.5[C or history of fever within the last 48 hours, other non-specific symptoms of acute febrile illness (suspected malaria), and written consent was obtained, including from parents/guardians if less than 18 years of age. Demographic, clinical, and epidemiological participant data were collected using a standardised electronic case record form (REDCap, version 10.6) by trained hospital staff.

### Blood sampling

Venous whole blood samples (or a capillary sample in those 10 years old or younger), were collected in ethylenediaminetetraacetic acid (EDTA) for routine hospital laboratory-based haematology. A 300μl whole blood aliquot was placed in field-stable DNA/RNA Shield^TM^ (Zymo Research, USA) total nucleic acid preservation media and frozen at -80°C prior to shipping with cold-chain maintenance to the study research laboratory in Jakarta.

### Health facility malaria screening

Standard local Ministry of Health point-of-care malaria testing was conducted at health facilities on all patients, consisting of a lateral-flow malaria rapid diagnostic test (RDT) with *P. falciparum*-specific histidine-rich-protein-2 (Pf-HRP2) and non-specific *Plasmodium* species parasite lactate dehydrogenase (PAN-pLDH) targets (CareStart Pf/PAN^TM^; Access Bio, USA). Giemsa blood films were read by local health facility microscopists blinded to the RDT result. *Plasmodium* species identification and parasite count quantification was conducted according to standard WHO microscopy procedures^36^.

### *Plasmodium* species PCR detection in human samples

*Plasmodium* genus real-time PCR targeting 18S ssu rRNA^37,38^ was performed with a QuantiTect Probe RT-PCR Kit following extraction of total nucleic acids (QIAamp® DNA Blood Mini Kit) from 200μL of mixed whole blood in DNA/RNA Shield and high-capacity cDNA reverse transcription (Applied Biosystems, USA). Positive results were defined as Ct values of less than 40 from both duplicate runs. Those positive had species-specific PCR assays conducted on the same cDNA samples including *Plasmodium* genus amplification^39^ followed by nested *P. knowlesi*^40^, *P. cynomolgi*^41^ and other human *Plasmodium* species (*P. falciparum, P. vivax, P. ovale spp.,* and *P. malariae*^39^) detection. Amplified nested PCR products were separated by electrophoresis using a 2% agarose gel stained by SYBR Safe™ (Invitrogen) and visualised on a UV transilluminator. Each PCR amplification included a *Plasmodium* species positive and negative control and molecular weight standards (Applied Biosystems, USA).

### Entomological case investigation and Ministry of Health survey data from Kalimantan

Patients with *P. knowlesi*-confirmed infections from North Kalimantan were interviewed by entomology team members. Standardised malaria case data collection included where the participant lived or worked up to 4 weeks prior to health facility presentation (encompassing the pre-patent period of *P. knowlesi* infection^5^). High-risk locations of *P. knowlesi* transmission were selected for spot entomology surveys to determine the presence of *Anopheles leucosphyrus* group and other *Anopheles* species with zoonotic malaria vector potential. Adult mosquito collections were conducted for a single night using hourly outdoor human-landing collections (HLC), all night from 6pm to 6am. There were six outdoor HLC collectors for each night and study site. These were complemented by *Anopheles* mosquito abundance data from harmonised entomology single night spot survey methods (6pm to 6am) conducted by the Ministry of Health, Indonesia and institutional partners as part of a national-scale disease vector and reservoir research program (*Rikhus Vektora*) from 2016 to 2018^42^. These latter surveys were carried out across multiple districts and classified environments (village, farm, forest, forest-edge, coastal mangrove/lagoon, coastal village) in North, Central, South and West Kalimantan provinces. Mosquito trapping methods included HLC (indoor and outdoor), animal-landing catches (cow or goat), animal-baited traps (single cow in tent), and CDC light traps (2 traps per habitat with octanol attractant).

### Mosquito morphological identification and *Plasmodium* species PCR detection

All mosquitoes were killed using chloroform. *Anopheles* species were separated and morphologically identified under a stereomicroscope to *Anopheles* genus^43^ and *Anopheles leucosphyrus* group species^27^. Mosquito abdomens were removed, and heads and thoraxes placed in perforated 1.5µL microcentrifuge tubes with silica gel prior to being individually homogenized using a pellet pestle in 100µl phosphate-buffered saline (PBS). DNA extraction was performed using a Quick-DNA Miniprep Plus Kit (Zymo Research, USA). Subsequently, a harmonised workflow was conducted to detect individual *Plasmodium* species 18S ssu rRNA gene targets as detailed above, but without reverse transcription. Additional testing for *P. inui*, *P. fieldi*, and *P. coatneyi* was also performed^41^.

### Statistical analysis

All statistical analyses were performed using R studio (version 2024.04.02). Chi-squared or Fisher’s exact test evaluated proportional differences in binary clinical and epidemiological variables between groups; Student’s t-test or Wilcoxon ranksum were used for pairwise comparisons of normal and non-normally distributed data, respectively. Results of microscopy and RDT assays evaluated against reference PCR were defined as true positive (TP), false negative (FN), true negative (TN), and false positive (FP), enabling calculation of diagnostic sensitivity (TP/TP+FN) and specificity (TN/TN+FP) with exact binomial 95% confidence intervals. The annual parasite index was calculated as the number of malaria positive cases divided by the number tested per 1000 at-risk people per year. Study site district catchment populations were estimated from 2020 census data projections^44^. Mosquito abundance data were analyzed using a generalized linear mixed-effects model (GLMMs) with a negative binomial distribution^45^. The initial model included *Anopheles* mosquito group (*leucosphyrus*, *barbirostris*, other), land type, mosquito collection method, and their interaction as fixed-effects, with random intercepts for district, province, and month to control for spatial and temporal variation. Models were compared using Akaike Information Criterion (AIC). A zero-inflation parameter was included and tested via residual diagnostics^46^. Estimated marginal means (EMMs) and pairwise contrasts were calculated^47^, with Tukey adjustments for multiple comparisons. Mosquito species diversity was calculated using the Shannon-Weiner Diversity Index (H) at the district and land type level^48^.

### Ethical approval

Ethical approval for the human surveillance was obtained from the research ethics commission of the Eijkman Institute for Molecular Biology (EIREC #139-2019). Permission to use entomology data was approved by the National Institute of Health Research and Development, Ministry of Health of Indonesia (Ref. No. TU:14/2/2020)

## Results

### Study enrolments and malaria case incidence

From December 2019 to September 2020 there were 429 patients with acute febrile illness enrolled in the study: 81 patients from Malinau and 348 from Sabang (**Table 1**). Overall, there were 15 patients (18.5%; 95%CI 10.8-28.7%) positive for malaria from Malinau diagnosed using PCR, comprising 10 *P. knowlesi* and 5 *P. vivax* infections. *Plasmodium knowlesi* infections represented 12.3 (95%CI 6.1-21.5%) of those in Malinau enrolled with fever. Sabang had 6 positive malaria cases: 4 *P. knowlesi* infections and 2 *Plasmodium* genus positive (species not identified). *Plasmodium knowlesi* cases comprised 1.1% (95%CI 0.3% to 2.9%) of the febrile patients enrolled. The *P. knowlesi* annual parasite index was 122 for 1000 people at-risk per year in Malinau and 11.5 in Sabang. The *P. knowlesi* crude incidence was 47.9 and 23.3 cases per 100,000 at-risk people each year for Malinau and Sabang districts, respectively. *Plasmodium cynomolgi* infections were not detected.

**Table 1.** clinical and laboratory features of *Plasmodium* species infections.

| Patient characteristic | North Kalimantan |  |  |  | Sabang |  |  |
| --- | --- | --- | --- | --- | --- | --- | --- |
|  | *P. knowlesi* | *P. vivax* | Controls | *P-value*<br>(*Pk* vs control) | *P. knowlesi* | Controls | *P-value*<br>(*Pk* vs control) |
| PCR confirmation, n (%) [95% CI] | 10<br>(12.2)<br>[6.0-21.3] | 5<br>(6.0)<br>[2.0-13.7] | 66<br>(81.5)<br>[71.3-89.2] | - | 4<br>(1.1)<br>[0.3-2.9] | 342<br>(98.8)<br>[97.1-99.7] | - |
| Age, years |  |  |  |  |  |  |  |
| Median | 41 | 36 | 37 | 0.632 | 35 | 24 | 0.220 |
| IQR | 41-42 | 29-54 | 27-52 |  | 26-48 | 10-40 |  |
| Range | 22-58 | 22-54 | 5-71 |  | 19-59 | 1-81 |  |
| Children (age <15 yrs), n (%) | 0 (0) | 0 (0) | 5 (9) | 0.999 | 0 (0) | 120 (35) | 0.302 |
| Male gender, n (%) | 10 (100) | 5 (100) | 44 (67) | **0.030** | 4 (100) | 183 (54) | 0.064 |
| Previous malaria (self-reported), n (%) | 3 (30) | 3 (60) | 13 (20) | 0.443 | 3 (75) | 20 (6) | **0.001** |
| Symptoms on enrolment, n (%) |  |  |  |  |  |  |  |
| Headache | 7 (70) | 5 (100) | 56 (88) | 0.427 | 4 (100) | 242 (71) | 0.601 |
| Nausea | 8 (80) | 3 (60) | 30 (48) | 0.088 | 2 (50) | 147 (43) | 0.999 |
| Cough | 6 (60) | 2 (40) | 27 (43) | 0.133 | 3 (75) | 168 (49) | 0.368 |
| Shortness of breath | 3 (33) | 1 (20) | 15 (25) | 0.687 | 0 (0) | 20 (6) | 0.617 |
| Vomiting | 5 (50) | 2 (20) | 17 (27) | 0.098 | 1 (25) | 68 (20) | 0.999 |
| Abdominal pain | 5 (50) | 1 (20) | 27 (44) | 0.130 | 1 (25) | 134 (39) | 0.490 |
| Fever (temperature >37.5°C) | 6 (60) | 4 (80) | 36 (55) | 0.999 | 2 (50) | 94 (28) | 0.311 |
| Febrile days (prior to presentation), n (IQR)[range] | 6<br>(5-6)[3-7] | 4<br>(3-4)[3-8] | 4<br>(3-6)[1-30] | 0.209 | 7<br>(5-7)[4-7] | 3<br>(1-4)[0-30] | **0.010** |
| Microscopy result <sup>†</sup> |  |  |  |  |  |  |  |
| *P. knowlesi* | 9 | 2 | 0 | - | 4 | 0 | - |
| *P. vivax* | 0 | 2 | 1 | - | 0 | 0 | - |
| *P. falciparum* | 1 | 0 | 0 | - | 0 | 0 | - |
| Negative | 0 | 0 | 66 | - | 0 | 342 | - |
| Sensitivity, % (95%CI) | 90.0 (55.5-99.7) | 50.0 (6.8-93.2) | - | - | 100 (39.8-100) | - | - |
| Specificity, % (95%CI) | 97.2 (90.2-99.7) | 98.7 (93.1-100) | - | - | 100 (98.9-100) | - | - |
| Parasite count |  |  |  |  |  |  |  |
| Slides read, n (%) | 10 (100) | 5 (100) | 67 (100) | - | 4 (100) | 344 (100) | - |
| Parasites/μL, geom. mean | 6,993 | 1,833 | - | - | 2,850 | - | - |
| Parasites/μL, median | 3,584 | 1,648 | - | - | 4,497 | - | - |
| IQR | 1,680-45,000 | 608-6,288 | - | - | 1,271-10,929 | - | - |
| Range | 512-180,000 | 512-6,416 | - | - | 287-15,120 | - | - |
| Parasites >15,000/μL, n (%) | 4 (40) | 0 (0) | 0.231 | - | 1 (25) | - | - |
| Parasites >100,000/μL, n (%) | 2 (20) | 0 (0) | 0.524 | - | 0 (0) | - | - |
| RDT result<sup>†</sup> |  |  |  |  |  |  |  |
| Pf-HRP2 positive | 0 | 0 | 0 | - | 1 | 0 | - |
| PAN pLDH positive | 10 | 5 | 0 | - | 3 | 0 | - |
| Sensitivity | 100 (69.2-100) | 100 (47.8-100) | - | - | 75.0 (19.4-99.4) | - | - |
| Specificity | 93.0 (84.3-97.7) | 86.8 (77.1-93.5) | - | - | 100 (98.9-100) | - | - |
| Anaemia* (baseline), n/N (%) | 0 (0) | 0 (0) | 0 (0) |  | 0 (0) | 1 (<1) |  |
| Platelet count, ×10<sup>3</sup>/μL, median (IQR) | 35 (18-46) | 66 (59-70) | 214 (148-287) | <0.001 | 78 (60-94) | 229 (184-274) | 0.001 |
| Thrombocytopenia, n/N (%)<br>(Platelet count <150×10<sup>3</sup>/μL) | 9 (90) | 5 (100) | 17 (26) | <0.001 | 4 (100) | 42 (13) | <0.001 |
| Severe malaria**, n (%) [95%CI] | 2 (20) [2.5-55.6] | 0 (0) | - | - | 0 (0) | - | - |

Control = participant malaria-negative on PCR with acute febrile illness
<sup>†</sup> Sensitivity/specificity calculated for *P. knowlesi* mono-infection vs non-*P. knowlesi* detection on microscopy against reference PCR; or separately *P. vivax* vs non-*P. vivax*
<sup>‡</sup> Sensitivity/specificity calculated for PAN pLDH detection for *P. knowlesi* vs non-*P. knowlesi* on reference PCR; RDT not conducted on n=1 control from Malinau
\*Anaemia based on WHO 2011 haemoglobin measurement criteria
\*\*WHO selected criteria: only anaemia and hyperparasitaemia able to be evaluated on all patients; shock evaluated on 57% of patients; both severe *P. knowlesi* infected patients had hyperparasitaemia as sole criterion
There were no statistically significant differences in reported variables between *P. knowlesi* malaria cases from Nth Kalimantan compared to those from Sabang
Table does not include data from the n=2 *Plasmodium* genus (species not determined) infections in Sabang, Aceh

### Demographic and clinical features

Of the 14 *P. knowlesi* infections, there were no differences in demographic or clinical features at presentation between locations, with all being adult males (**Table 1**). In Malinau the median age for *P. knowlesi* malaria cases was 41 years, with a range of 22 to 58 years. Three patients at each site reported a previous health facility diagnosis of malaria. Overall, patients reported a median of 6 days of fever prior to presentation, similar to those negative for malaria. Those with *P. knowlesi* infections commonly presented with non-specific symptoms including reported headache, nausea and cough, with abdominal pain present in half of patients. Thrombocytopaenia was present in 90% of patients with malaria, compared to 14% overall in febrile controls (p<0.001)

### Microscopy and RDT diagnostics

A single *P. knowlesi* infection was misidentified as *P. falciparum*, and 2 *P. vivax* infections misidentified as *P. knowlesi* by routine microscopy at Malinau hospital, resulting in a diagnostic sensitivity of 90% (95% CI 55.5-99.7%) and specificity of 97.2% (95%CI 93.1-100%) (**Table 1**). In Sabang all 4 *P. knowlesi* infections were positively identified by microscopy, resulting in 100% sensitivity and specificity. The 2 undifferentiated *Plasmodium* genus infections were negative on microscopy and RDT. The PAN-pLDH RDT detected all 10 *P. knowlesi* and 5 *P. vivax* infections at Malinau. For Sabang a single *P. knowlesi* infection on the PAN-pLDH test strip was not detected despite a parasitaemia of 6,738/µL.

### Parasitaemia and severe disease

The geometric mean parasite count of *P. knowlesi* infections was higher in Malinau patients at 6,993 parasites/µL (range: 512 to 180,000/µL) compared to those from Sabang with a mean of 2,850 parasites/µL (range: 287 to 15,120/µL) (**Table 1**). Four patients from Malinau had parasite counts greater than 15,000/µL, including 2 (20%) patients with hyperparasitaemia (parasite counts of 155,000/µL and 180,000/µL), defined as severe malaria by WHO research criteria^49^. Other severe malaria manifestations were not recorded: blood pressure was only recorded in 57% of patients, and other severe malaria criteria including acute kidney injury, acute respiratory distress, hyperbilirubinaemia, hypoglycaemia and metabolic acidosis were unable to be assessed due to a lack of hospital laboratory capacity. The patients with severe knowlesi malaria were both adult males, aged between 30 and 50 years of age. Both patients had a recent history of sleeping outside, had spent extended periods in intact forest areas (hunting and logging respectively), and presented after more than 5 days of fever, abdominal pain, headache, and vomiting. In Sabang, all 4 patients with *P. knowlesi* malaria had uncomplicated disease.

### Antimalarial treatment

In Sabang all 4 patients with *P. knowlesi* malaria received appropriate oral treatment with dihydroartemesinin-piperaquine, and in addition were given a single dose of primaquine. At Malinau Hospital, due to shortages of standard antimalarial drugs during the COVID-19 pandemic, one of the patients with severe *P. knowlesi* malaria received 3 days of oral quinine and doxycycline in addition to a single dose of primaquine. The majority of the remaining *P. knowlesi*- or *P. vivax*-infected patients did not have their specific blood-stage antimalarial treatment recorded. Although no deaths were reported, there was no formal follow-up for clinical outcomes as part of this pandemic-limited observational study.

### Epidemiology

In Malinau, patients with *P. knowlesi* infections included 3 identifying as farmers, in addition to an individual forestry worker, builder/carpenter, teacher, head of village, public servant, driver, and phone tower technician (**Supplemental Table 1**). For Sabang, knowlesi malaria cases included two patients self-employed/small business owners and a single student. There were no plantation workers among malaria cases, and only 2 (3%) documented in the Malinau control group. In comparison to febrile controls, the majority of those with *P. knowlesi* malaria reported a recent travel history requiring sleeping outside of their usual household in the last 2 weeks, including 80% of *P. knowlesi* cases in Malinau (p<0.001), and 50% in Sabang (p=0.021). Antimalarial bed net use was low overall and not associated with protection from malaria, with 26% of patients recently sleeping under a bednet (of which 3% were insecticide treated). Awareness of monkey presence was reported in 50% and 100% of patients with knowlesi malaria from Malinau and Sabang, respectively. A higher proportion of *P. knowlesi* malaria cases from Malinau reported spending time in intact forest areas in the preceding 2 weeks compared to febrile controls (80% vs 30% respectively; OR 9.3 [95%CI 1.8-47.8]; p=0.004). None of the four patients with *P. knowlesi* infections in Sabang had a recent history of forest exposure. In Malinau 30% of *P. knowlesi* malaria cases reported charcoal making as their primary forest-related activity compared to 5% of controls (p=0.027). Household construction or surrounding environmental features were not associated with *P. knowlesi* infections.

**Supplemental Table 1.** Epidemiological factors associated with *P. knowlesi* infections.

| Patient characteristic | North Kalimantan |  |  |  | Sabang |  |  |
| --- | --- | --- | --- | --- | --- | --- | --- |
|  | *P. knowlesi*<br>n=10 | *P. vivax*<br>n=5 | Controls<br>n=67 | *P-value*<br>(*Pk* vs<br>control) | *P. knowlesi*<br>n=4 | Controls<br>n=342 | *P-value*<br>(*Pk* vs<br>control) |
| **Occupation** |  |  |  |  |  |  |  |
| Farmer | 3 (30) | 0 | 15 (23) | 0.448 | 0 | 19 (6) | 0.794 |
| Plantation work | 0 | 0 | 2 (3) | 0.750 | 0 | 1 (<1) | 0.988 |
| Forestry | 1 (10) | 0 | 2 (3) | 0.353 | 0 | 1 (<1) | 0.988 |
| Builder/carpenter | 1 (10) | 0 | 0 | 0.133 | 0 | 4 (1) | 0.954 |
| Student | 0 | 0 | 7 (11) | 0.351 | 1 (25) | 119 (35) | 0.558 |
| Business/self-employed | 0 | 2 (20) | 3 (5) | 0.647 | 2 (25) | 24 (7) | **0.031** |
| Household/family carer | 0 | 0 | 5 (8) | 0.479 | 0 | 56 (17) | 0.485 |
| Public servant | 2 (20) | 2 (20) | 10 (15) | 0.503 | 0 | 19 (6) | 0.794 |
| Fisherman | 0 | 0 | 0 | - | 0 | 14 (4) | 0.845 |
| Unemployed | 0 | 0 | 1 (2) | 0.867 | 1 (25) | 34 (10) | 0.345 |
| Other | 3 (30) | 1 (20) | 20 (31) | 0.637 | 0 | 45 (13) | 0.565 |
| **Activities** |  |  |  |  |  |  |  |
| Sleep outside house (last 2 weeks) | 8 (80) | 2 (40) | 12 (18) | **<0.001** | 2 (50) | 20 (6) | **0.021** |
| Bednet use outside house | 1 (14) | 0 | 0 | 0.101 | 0 | 1 (<1) | 0.988 |
| Aware of monkeys | 5 (50) | 1 (20) | 14 (21) | 0.064 | 4 (100) | 143 (42) | **0.032** |
| Clearing forest/vegetation | 5 (50) | 2 (40) | 27 (42) | 0.432 | 0 | 43 (13) | 0.586 |
| Forest exposure (>4 hours) | 8 (80) | 3 (60) | 19 (30) | **0.004** | 0 | 29 (9) | 0.702 |
| Cutting timber | 1 (10) | 1 (20) | 0 | 0.132 | 0 | 3 (<1) | 0.966 |
| Collecting wood | 0 | 0 | 0 | - | 0 | 2 (<1) | 0.977 |
| Hunting | 2 (20) | 1 (20) | 3 (5) | 0.126 | 0 | 1 (<1) | 0.988 |
| Charcoal making | 3 (30) | 0 | 3 (5) | **0.027** | 0 | 2 (<1) | 0.977 |
| Other forest activity | 3 (30) | 2 (40) | 14 (21) | 0.684 | 0 | 22 (6) | 0.999 |
| **Malaria prevention** |  |  |  |  |  |  |  |
| Bed net use (any) | 3 (30) | 1 (20) | 21 (32) | 0.610 | 2 (50) | 85 (25) | 0.264 |
| Bed net use (Insecticide treated) | 0 | 0 | 3 (5) | 0.611 | 1 (25) | 51 (15) | 0.482 |
| Personal insect repellent use<br>(lotion/spray) | 4 (40) | 2 (40) | 16 (25) | 0.262 | 0 | 43 (13) | 0.587 |
| **Household** |  |  |  |  |  |  |  |
| Wall construction |  |  |  |  |  |  |  |
| Wood | 8 (89) | 5 (100) | 56 (85) | 0.607 | 1 (25) | 72 (21) | 0.614 |
| Concrete | 1 (11) | 0 | 7 (11) | 0.660 | 2 (50) | 252 (74) | 0.288 |
| Other | 0 | 0 | 3 (5) | 0.678 | 1 (25) | 18 (5) | 0.203 |
| Elevated (>1m stilts) | 7 (70) | 3 (60) | 55 (85) | 0.234 | 4 (100) | 231 (68) | 0.211 |
| Open eaves/gaps | 8 (80) | 3 (75) | 48 (74) | 0.510 | 3 (75) | 149 (44) | 0.226 |
| Insecticide treated walls (IRS) | 1 (13) | 0 | 2 (3) | 0.321 | 1 (25) | 71 (21) | 0.612 |
| ***Household environment (±100m)*** |  |  |  |  |  |  |  |
| Oil palm | 0 | 0 | 4 (6) | 0.618 | 0 | 0 | - |
| Rubber | 0 | 0 | 0 | - | 0 | 1 (<1) | 0.988 |
| Rice paddy | 0 | 1 (20) | 9 (14) | 0.260 | 0 | 0 | - |
| Mangrove | 1 (10) | 0 | 1 (1) | 0.250 | 0 | 6 (2) | 0.932 |
| Cleared forest area | 2 (20) | 0 | 6 (9) | 0.288 | 1 (25) | 26 (8) | 0.279 |
| Intact forest area | 3 (30) | 2 (40) | 19 (30) | 0.623 | 3 (75) | 121 (35) | 0.133 |

The difference in forest exposure (>4 h) between *P. knowlesi* malaria cases in Nth Kalimantan was higher compared to those from Sabang (p=0.015); there were no other statistically significant differences within those with *P. knowlesi* infections across sites.

### Entomological malaria case investigation in North Kalimantan

Field mosquito collections were carried out overnight using HLC in 5 separate study sites from May to August 2022, including: Malinau (a residential area), Pulau Sapi (rubber plantation/mixed garden), Long Tarik (rainforest with a swamp), Long Metut (village bordering forest) and Jelukut (rainforest). There were 7 forestry workers with a history of travel to Jelukut who reported staying for about one month before developing fever and testing positive for *P. knowlesi* infections. Pig-tailed macaques (*M. nemestrina*) were reported to be frequently present in this area. Eight *Anopheles* species comprising 116 mosquitos were captured (**Figure 2**), with the majority (76%) identified as *An. barbirostris* of which nearly all were captured at Pulau Sapi. A single specimen of *An. balabacensis* was collected in Jelukut (at 7pm), in addition to several *Anopheles* larvae. All *Anopheles* samples were negative for *Plasmodium* on PCR.

**Figure 2.**
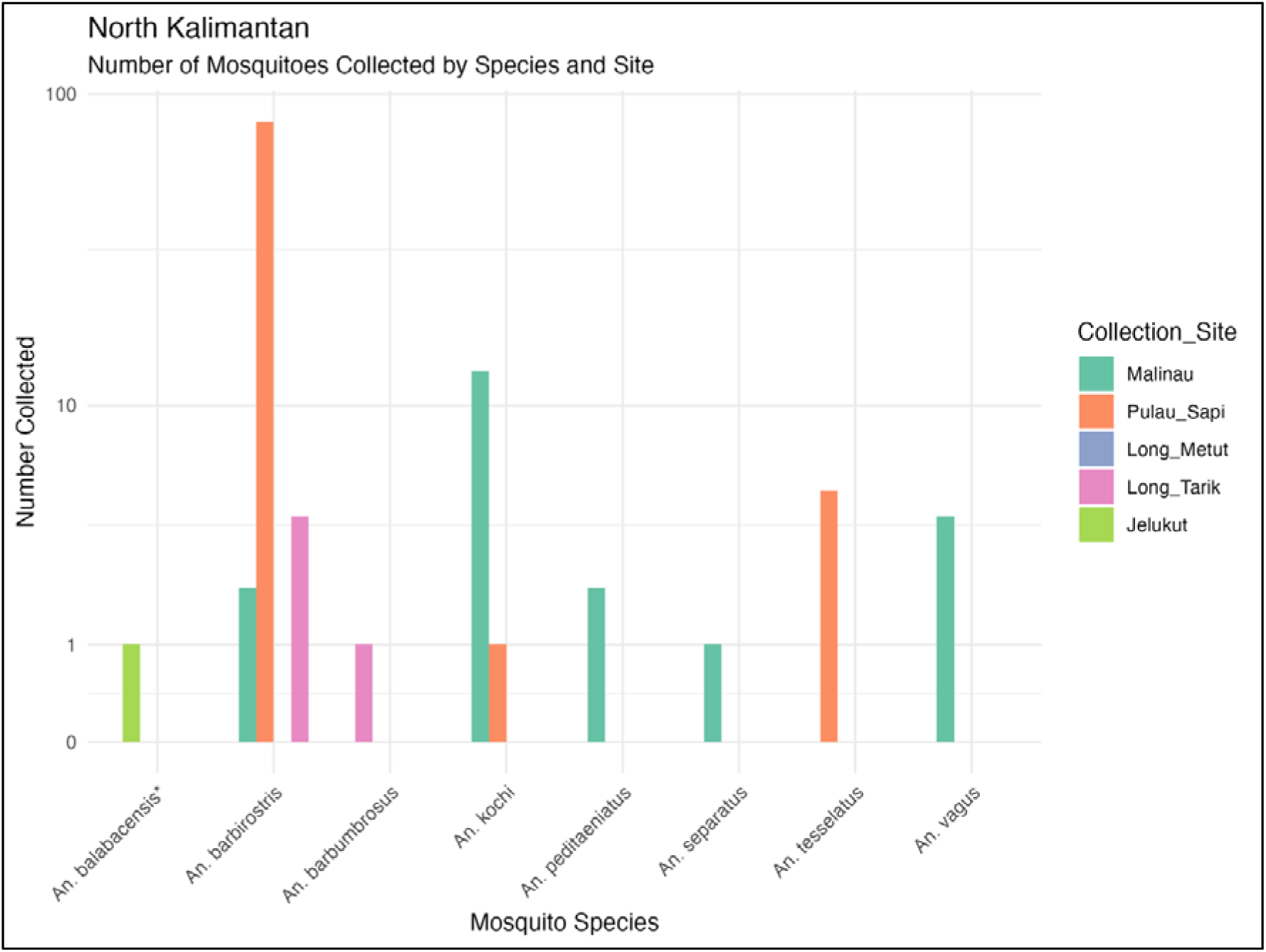
*Anopheles* species presence at locations with *P. knowlesi* human infection index cases in Malinau, North Kalimantan

### Entomological surveillance data across four Kalimantan provinces

Mosquito occurrence data were re-analysed from overnight collections in North Kalimantan (July to August 2018), Central Kalimantan (May 2017) and South and West Kalimantan (July to August 2016). At least 32 *Anopheles* mosquito species were morphologically identified across different land types in Kalimantan (**Figure 3**). In North Kalimantan, there were 18 *Anopheles* species, including *An. balabacensis* (only found in Nunakan district in North Kalimantan) and *An. latens* (also captured in Central Kalimantan). Central Kalimantan had the highest diversity of *Anopheles leucosphyrus* group mosquitos with 4 species identified, including the suspected zoonotic malaria vectors *An. baimaii* and *An. leucosphyrus*, and the primarily zoophilic *An. hackeri. Anopheles leucosphyrus* was the only potential zoonotic vector species found in South Kalimantan. All *Anopheles* samples were PCR-negative for *Plasmodium* species^42^.

**Figure 3:**
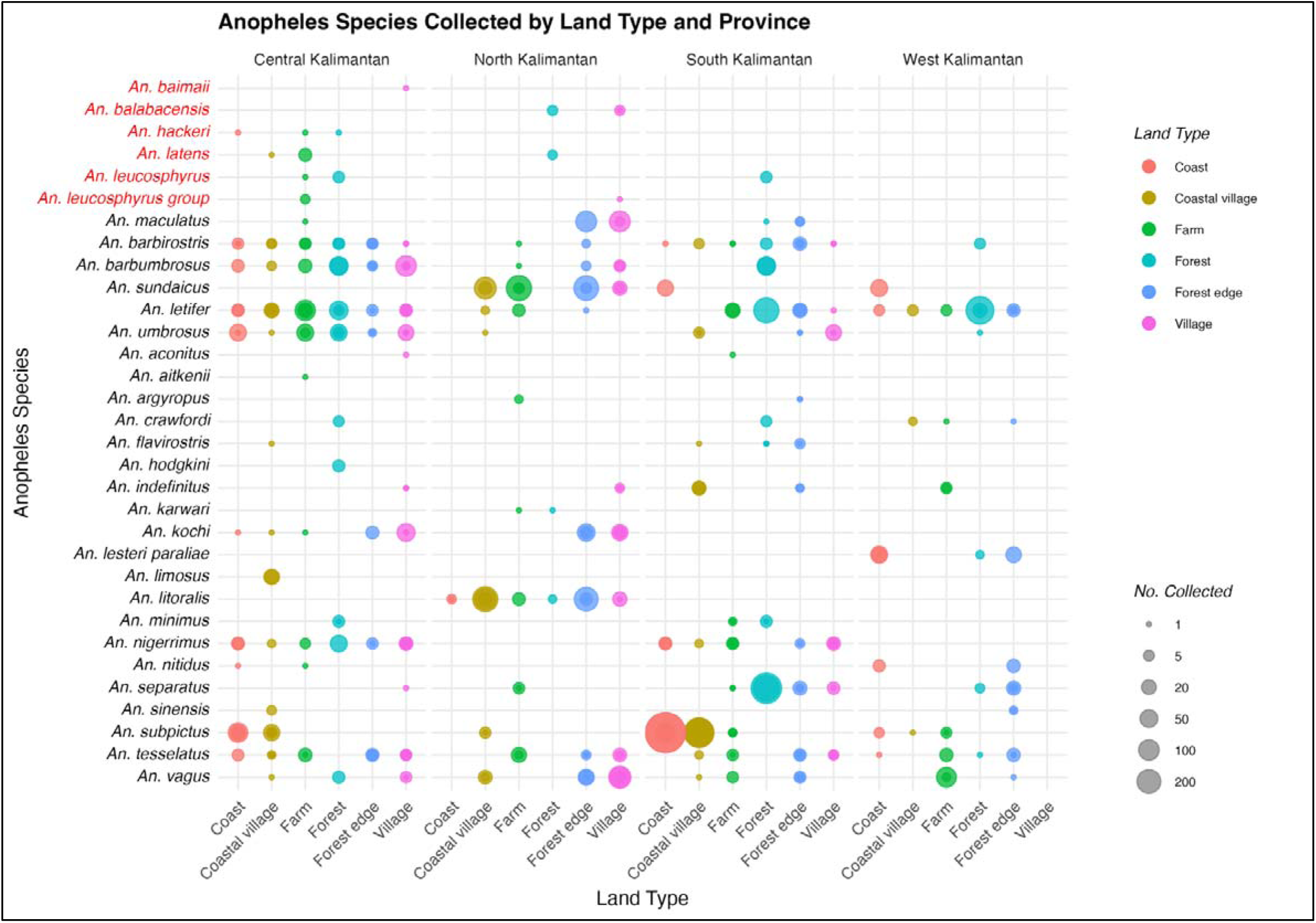
Presence of *Anopheles* mosquito species collected across different land types in four provinces in Kalimantan 2016-18 (circle sizes represent square-root scaled counts per species, with Leucosphyrus group species labelled in red)

*Anopheles leucosphyrus* group mosquitos were found in forest (45%), farm (36%), and village (36%) habitats, and for *An. hackeri* and *An. latens* also in coastal mangrove/lagoon and coastal village sites, respectively. GLMM model results predicted significantly lower relative abundances of Leucosphyrus group (5%) and Barbirostris group (25%) mosquitoes in coastal land types (mangrove area and village) compared to other *Anopheles* species (p ≤ 0.037), with no significant differences observed across farm, forest, or village habitats. *Anopheles* species diversity overall measured by Shannon index across districts was highest in farm areas (H′ = 0.84, SD = 0.49) and lowest in coastal mangrove areas (H′ = 0.56, SD = 0.52), although differences were not statistically significant.

Across Kalimantan, outdoor HLC was the most commonly deployed and effective collection method across all provinces and land types for *An. leucosphyrus* group mosquitos (**Figure 4**). However, a single *An. balabacensis* mosquito was captured indoors using HLC, and an *An. leucosphyrus* group mosquito (species undetermined) was captured using an animal-baited trap in a village site. Post-hoc pairwise comparisons using only outdoor HLC collection data across land types showed no significant differences in Leucosphyrus group abundance, with the highest predicted abundance similar in farm and forest sites, and reductions of ∼60% and 75% in village and coastal sites, respectively.

**Figure 4:**
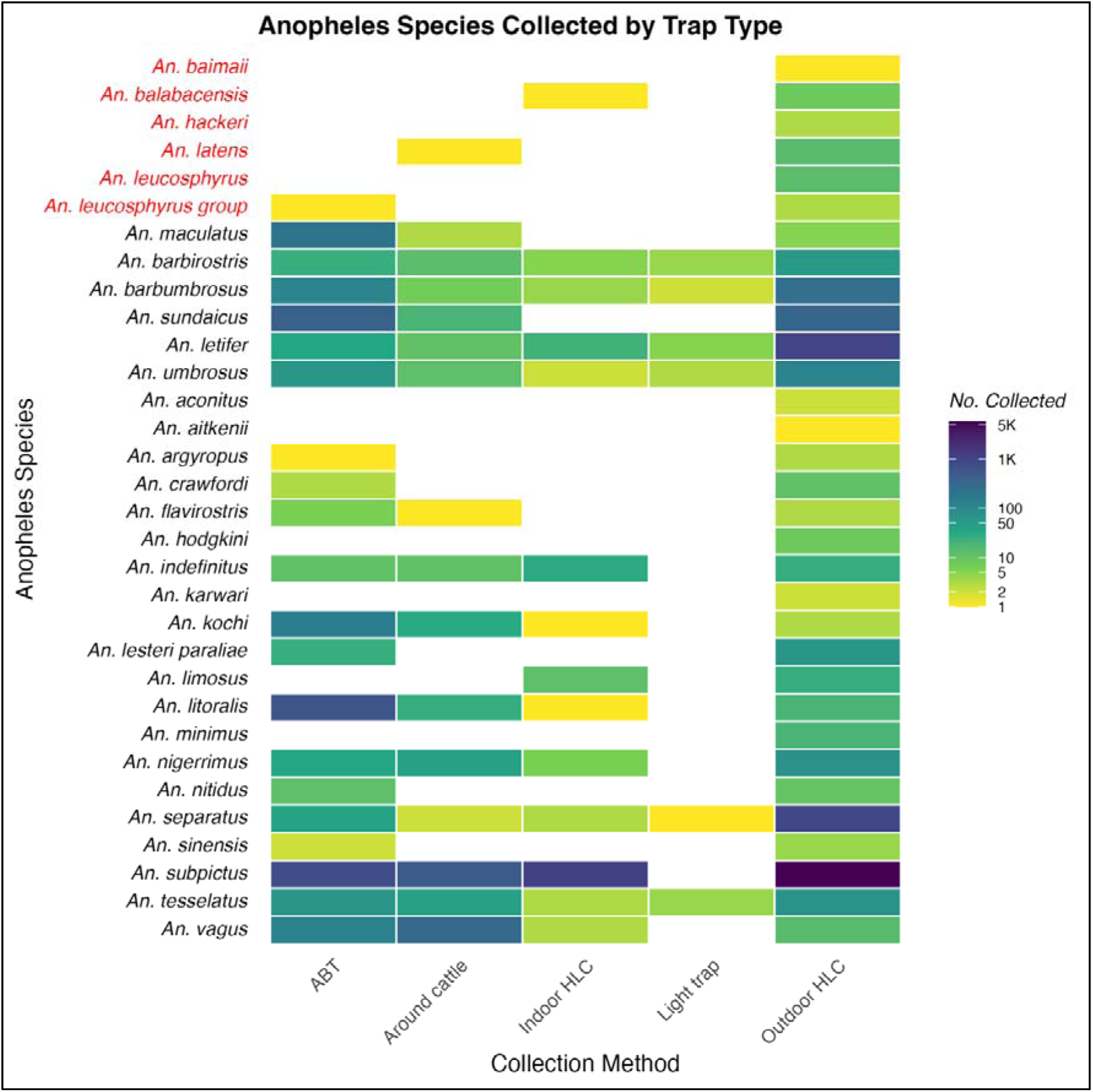
Comparison of *Anopheles* species collected using different traps across Kalimantan provinces 2016-18 (Number collected represents mosquito abundance in log_10_ scale; Leucosphyrus group species are labelled in red)

## Discussion

This study confirms zoonotic *P. knowlesi* as the major cause of malaria in North Kalimantan and Sabang, Aceh, through molecular confirmation of symptomatic infections in patients presenting to district-level health facilities. The presence of previously incriminated *P. knowlesi* vectors *An. balabacensis*^25^ and *An. latens*^29^ was also confirmed in Kalimantan. We report confirmed WHO-defined severe malaria^49^ due to *P. knowlesi* in North Kalimantan. These findings suggest *P. knowlesi* malaria may contribute substantially to the disease burden of non-specific acute febrile illness in remote populations in Kalimantan, consistent with neighbouring regions of Malaysia^50^. The presence of *P. knowlesi* infections across both North Kalimantan and the geographically remote island of Sabang, Aceh, underscores the importance of ongoing access to appropriate malaria diagnostics– including routine point-of-care tools and molecular surveillance^51^ –and timely availability of national antimalarial treatment at regional health facilities in areas of zoonotic malaria risk. Sustained malaria testing remains critical, even in provinces nearing elimination of non-zoonotic *Plasmodium* as part of Indonesia’s subnational WHO malaria elimination goals^52^. Spatial understanding of zoonotic malaria transmission was enhanced through analysis of entomological case investigation and Ministry of Health surveillance data. Species-specific differences in *Anopheles leucosphyrus* group distributions were observed across provinces and land types. Importantly, we documented the widespread presence of predominantly forest-associated *Anopheles* zoonotic malaria vectors in farm and village environments, reflecting their adaptation and expanded ecological range^31^.

The passive health facility surveillance design for this study employed routine point-of-care diagnostics with molecular confirmation of suspected malaria cases, revealing previously undocumented *P. knowlesi* transmission as the most common cause of malaria in Malinau, North Kalimantan. A higher proportion of *P. knowlesi*-positive cases among febrile presentations occurred in Malinau (12.5%) compared to Sabang, Aceh (1.5%) where vector species^53^ macaque host presence^54^ and ecological risks differ^18,19^. In Sabang, *P. knowlesi* was the only cause of malaria detected, consistent with reported progress towards elimination of non-zoonotic *Plasmodium*^55^. All confirmed *P. knowlesi* cases presented with non-specific symptoms, reflecting overlap with other febrile illnesses and under-detection if malaria is not clinically suspected. Despite sampling limitations due to the small number of patients enrolled in Malinau during the COVID-19 pandemic, district case incidence was higher than the overall Ministry of Health reported malaria trends in Kalimantan^56^, suggesting an undetected zoonotic malaria burden or localised high-transmission areas. Malaria notifications in Kalimantan have remained low since at least 2014, with an annual parasite index below 1 per 1000 people^56^. In 2023, only 145 malaria cases were reported across North Kalimantan, including 19 from Malinau, with a slight provincial-level incidence increase from 0.3 to 0.4 per 1000 people since 2019. In neighbouring East Kalimantan, cases rose from ∼2000 in previous years to over 3000 in 2023, coinciding with a worker influx and deforestation linked to the construction of the new national capital^20^. However, few PCR-confirmed *P. knowlesi* cases have been reported to date from East Kalimantan^12^. In comparison, 16 malaria cases (species not reported) were recorded from passive surveillance in Sabang, Aceh (population 42,888) in 2023^56^.

Point-of-care diagnostics performed well for detecting patent *P. knowlesi* infections, with microscopy sensitivity over 90% and specificity above 97%—higher than expected based on previous data from Aceh^5^ and neighbouring Malaysia^57^. All cases were initially enrolled as suspected malaria based on clinical presentation, with microscopy findings highlighting increased awareness of zoonotic malaria by health facility microscopists and the near-absence of *P. falciparum* and *P. vivax* in the region. There are clinical implications for the small number of *P. vivax* infections misidentified as *P. knowlesi* using point-of-care tools, where lack of timely molecular confirmation means primaquine for radical cure may not be provided. Pan-pLDH RDTs detected *P. knowlesi* in most cases, with the lowest positive result at 287 parasites/μL. A single high-density *P. knowlesi*-infected case (6,738/μL) tested negative, consistent with reports of improving sensitivity of newer RDTs above 200 parasites/µL, despite variable performance even at moderate parasitaemia and possible reader error^21^. These findings underscore both the utility and limitations of current diagnostic tools to detect *P. knowlesi* in low-endemic settings, where clinical suspicion often prompts testing rather than mandated febrile illness surveillance. Standard RDTs targeting *P. falciparum*-HRP2 and pan-pLDH cannot differentiate non-*P. falciparum* species^23,24^. At the time of the study, only ∼2000 point-of-care malaria tests were conducted annually across 52 health facilities in North Kalimantan^56^, with Malinau comprising only a small portion despite a high proportion of *P. knowlesi* cases.

The confirmation of WHO-defined severe malaria caused by *P. knowlesi* infection in North Kalimantan adds to growing regional evidence that severe disease may occur wherever *P. knowlesi* transmission exists^58–61^. Whether a greater parasite inoculation in higher-risk populations in high transmission areas contributes to the increased parasitemia and risk of severe disease seen in these groups, as suggested by Malaysian data^62,63^, remains speculative. While most reports of severe morbidity are concentrated in high-transmission areas of Malaysian Borneo, the role of other potential protective or disease-enhancing factors—such as cross-protective immunity^64^, G6PD deficiency^65^, or parasite genetic determinants of red blood cell invasion^66,67^—remains unclear in shaping geographical distribution patterns and risk. Severe *P. knowlesi* malaria has been previously reported elsewhere in Kalimantan, including patients in Central Kalimantan with hyperparasitaemia (∼215,000 parasites/μL)^11^, and in East Kalimantan with severe anaemia (haemoglobin 5.3 g/dL) and decreased consciousness^68^. In our study, we were unable to assess the full range of WHO-defined severity research criteria, including acute kidney injury, jaundice and metabolic acidosis, due to limited hospital laboratory capacity for biochemical testing, despite these complications being commonly reported^58,59,63^. Microscopic quantification of parasitaemia remains a valuable tool for risk assessment in the absence of other laboratory capacity, with >15,000 parasites/µL associated with a 16-fold increased risk and 98% negative predictive value for severe complications^59^. Accurate diagnosis and early risk assessment are critical to enable prompt administration of intravenous artesunate, which reduces fatalities^63,69^. Although Indonesian guidelines recommend intravenous artesunate followed by oral artemisinin-based combination therapy for severe malaria caused by any *Plasmodium* species^70^, implementation remains challenging in remote settings where access to essential antimalarials can be limited. These challenges are further compounded by reduced clinical suspicion in areas nearing elimination of non-zoonotic *Plasmodium* species.

A wide range of occupations and forest-related activities were reported among patients with *P. knowlesi* malaria, including farming and hunting, reflecting diverse behavioural risk profiles. Forest activities remain a key driver of *P. knowlesi* transmission^71^, with positive associations with sleeping outdoors, and awareness of potential monkey reservoirs. Previously undocumented small-scale charcoal production was associated with higher zoonotic malaria risk, likely involving extended periods in forested areas harvesting wood and maintaining kilns. However, in contrast to neighbouring Malaysia^65^, plantation work was not reported as a primary occupation by any patient with *P. knowlesi* malaria and was rarely documented in malaria-negative controls. It is possible that large-scale oil palm plantation workers were under-represented, potentially due to self-identification as farmers, under-presentation to health facilities, or seeking treatment elsewhere. Patients with *P. knowlesi* malaria lacked associations with household construction or surrounding environmental features as reported in Sabah, Malaysia^65^. Despite the current analysis being constrained by low enrolment numbers, findings suggest infection was more likely acquired during time spent in remote forested settings rather than within the peri-domestic environment^65,71^. The absence of children among confirmed malaria cases, across all *Plasmodium* species, is consistent with the predominantly adult burden of disease in zoonotic transmission settings^59,72^ and areas approaching elimination of non-zoonotic *Plasmodium* species, reflecting both behavioural exposure and declining childhood-acquired immunity^73^.

Analysis of Ministry of Health entomological surveillance data across four major provinces in Kalimantan reinforces the widespread, although poorly characterised, heterogenous distribution of *Anopheles leucosphyrus* group species in the region^30,53,54^. *Anopheles balabacensis* and *An. latens*, both confirmed vectors of *P. knowlesi*^25,74,75^, were found in low numbers in North Kalimantan, and for the latter also in Central Kalimantan, highlighting the under-recognised risk of zoonotic malaria transmission south of Malaysian Borneo^19^. The ecological range of *An. leucosphyrus* group mosquitoes across forest, farm, and village habitats supports the need for surveillance strategies that include both human and vector components in forest-adjacent communities. However, no *An. leucosphyrus* group mosquitoes were reported from forest-edge habitats, suggesting possible gaps in sampling coverage or differences in habitat preference^76^. Although overall mosquito catches were low during single-night human landing catch (HLC) surveys, the detection of *An. balabacensis*^25,77,78^ and potential larval habitats for this mosquito species^26^ in the remote forest area of Jelukut where human cases were reported is epidemiologically significant. *Anopheles balabacensis* is widely distributed across different habitats including village locations, shrub-bush, forest-edge, and oil-palm plantations in neighbouring Malaysia^25,77^, with *P. knowlesi* co-infections found with other macaque parasite species *P. inui*, *P. coatneyi* and *P. cynomolgi* from mosquitos captured in the early evening^77^. Despite all *Anopheles* in this study testing PCR-negative for *P. knowlesi* and other *Plasmodium*, it is likely that *An. balabacensis* contributes significantly to zoonotic malaria transmission in this area of Indonesia. The absence of sporozoite-positive mosquitoes is not unexpected given the limited collections and typically low sporozoite rates in *An. leucosphyrus* group vectors even in high zoonotic transmission regions^31^.

The dominance of *An. barbirostris* group mosquitos in areas of Kalimantan highlights their ability to inhabit varied ecological niches and potentially contribute as secondary zoonotic vectors^79,80^, including in areas which remain receptive for non-zoonotic malaria. Although not identified in this study, the possible presence of *An. donaldi,* a member of the *An. barbirostris* group, could alter the distribution of at-risk populations, as previous studies in adjacent Malaysian areas have reported *P. knowlesi*-infected *An. donaldi* mosquitos by PCR^76,78^. Morphological differentiation of the closely related *An. barbirostris (s.l)* and *An. donaldi* is challenging, with molecular identification required to allow the correct classification of *P. knowlesi*-infected *An. donaldi* in Sarawak^78^. The reported presence of *An. baimaii*, a Leucosphyrus group member more commonly associated with the Greater Mekong Subregion would also benefit from definitive molecular identification^81^. In Sabang, Aceh, despite *P. knowlesi* infections in humans^4^, studies have yet to document *P. knowlesi* in *Anopheles* mosquitoes or long-tailed macaques^32,35^. Several *Anopheles* species have been identified, including *An. dirus,* and it is highly probable that both *An. dirus* and other members of the *An. leucosphyrus* group are significant vectors for local zoonotic malaria transmission. *Anopheles dirus* has previously been implicated in zoonotic malaria transmission across Southeast Asia^30,33,34^.

This study has several limitations. The small number of passively enrolled patients in Malinau, partly due to the COVID-19 pandemic, limits the generalisability of malaria incidence estimates. Future larger-scale molecular and integrated serological malaria surveillance^82,83^ would support more robust characterisation of population-level prevalence, disease burden and ecological risk associations. Underrepresentation of certain at-risk groups, such as plantation workers or children, may reflect health-seeking behaviour or access barriers and constrain interpretation of behavioural and demographic risk patterns. Molecular confirmation of *Plasmodium* species is not available in real-time, for which more sensitive and specific RDTs could improve treatment, particularly for *P. vivax*, and detect zoonotic malaria below microscopy or RDT thresholds^21^. Entomological Ministry of Health single night sampling was conducted at varying times and under differing environmental conditions, limiting interpretation of distribution and abundance estimates. More detailed classification of land types and longitudinal sampling design would provide greater information on vector behaviour and sporozoite-infection prevalence. Morphological misidentification of closely related *Anopheles* species, particularly within the Leucosphyrus and Barbirostris groups, cannot be ruled out due to a lack of molecular confirmation in the current study.

## Conclusion

This study confirms *P. knowlesi* as a common cause of non-specific febrile illness in North Kalimantan, and as a cause of severe malaria, with implications for diagnosis and treatment, including access, in low-transmission, elimination-phase settings. The widespread ecological range of competent vectors highlights the risk of zoonotic malaria transmission beyond traditional forest habitats. Continued access to appropriate diagnostics, strengthened molecular surveillance, and targeted vector monitoring are essential for effective zoonotic malaria control in Indonesia.

## Data Availability

All data produced in the present study are available upon reasonable request to the authors and with permission of Indonesian government agencies where relevant.

## Acknowledgements

We thank the study patients, the health facility directors and staff at the study sites, and all field mosquito collectors and all technical teams of the Rikhus Vektora project, and the Ministry of Health, Indonesia for the analysis of previously collected entomology data.

This work was supported by the Australian Centre for International Agricultural Research, Australian Government (LS-2018-214 and LS-2019-116), and the National Health and Medical Research Council, Australia (fellowships to NMA [1042072], MJG [1138860] and BEB [2016801]).

## Author contributions

RN, FNC, BEB, NMA, TAG, and MJG conceived the study design; LT, RA, PK, NF, FAN, TY, II, and H performed data collection and conducted the experiments; MJG, LT, RA, RN and TAG performed data analysis and wrote the original draft; NMA, BEB, TAG, RN and FNC provided supervision, funding acquisition and resources; All authors read, reviewed and approved the final manuscript.

